# Polygenic and transcriptional risk scores identify chronic obstructive pulmonary disease subtypes

**DOI:** 10.1101/2024.05.20.24307621

**Authors:** Matthew Moll, Julian Hecker, John Platig, Jingzhou Zhang, Auyon J. Ghosh, Katherine A. Pratte, Rui-Sheng Wang, Davin Hill, Iain R. Konigsberg, Joe W. Chiles, Craig P. Hersh, Peter J. Castaldi, Kimberly Glass, Jennifer G. Dy, Don D. Sin, Ruth Tal-Singer, Majd Mouded, Stephen I. Rennard, Gary P. Anderson, Gregory L. Kinney, Russell P. Bowler, Jeffrey L. Curtis, Merry-Lynn McDonald, Edwin K. Silverman, Brian D. Hobbs, Michael H. Cho

## Abstract

**Rationale:** Genetic variants and gene expression predict risk of chronic obstructive pulmonary disease (COPD), but their effect on COPD heterogeneity is unclear.

**Objectives:** Define high-risk COPD subtypes using both genetics (polygenic risk score, PRS) and blood gene expression (transcriptional risk score, TRS) and assess differences in clinical and molecular characteristics.

**Methods:** We defined high-risk groups based on PRS and TRS quantiles by maximizing differences in protein biomarkers in a COPDGene training set and identified these groups in COPDGene and ECLIPSE test sets. We tested multivariable associations of subgroups with clinical outcomes and compared protein-protein interaction networks and drug repurposing analyses between high-risk groups.

**Measurements and Main Results:** We examined two high-risk omics-defined groups in non-overlapping test sets (n=1,133 NHW COPDGene, n=299 African American (AA) COPDGene, n=468 ECLIPSE). We defined “High activity” (low PRS/high TRS) and “severe risk” (high PRS/high TRS) subgroups. Participants in both subgroups had lower body-mass index (BMI), lower lung function, and alterations in metabolic, growth, and immune signaling processes compared to a low-risk (low PRS, low TRS) reference subgroup. “High activity” but not “severe risk” participants had greater prospective FEV_1_ decline (COPDGene: -51 mL/year; ECLIPSE: - 40 mL/year) and their proteomic profiles were enriched in gene sets perturbed by treatment with 5-lipoxygenase inhibitors and angiotensin-converting enzyme (ACE) inhibitors.

**Conclusions:** Concomitant use of polygenic and transcriptional risk scores identified clinical and molecular heterogeneity amongst high-risk individuals. Proteomic and drug repurposing analysis identified subtype-specific enrichment for therapies and suggest prior drug repurposing failures may be explained by patient selection.

## INTRODUCTION

Chronic obstructive pulmonary disease (COPD) is a leading cause of morbidity and mortality worldwide^1^. Although COPD is characterized by irreversible airflow obstruction, there is marked heterogeneity amongst individuals in emphysema and airway pathology, exacerbation incidence, and lung function decline^2,3^. Identifying individuals at high risk for rapid COPD progression or eventual severe disease is critically important to implement personalized therapeutic approaches.

Large-scale omics data offer the potential to identify via a simple blood test high risk groups that share distinct, targetable pathobiology. Genetics, quantified with polygenic risk scores (PRSs), can identify individuals at high risk for coronary artery disease and guide consideration of statin therapy earlier than advised by current guidelines^4^. In cancer, integrating genetic and transcriptomic profiling can improve therapy recommendations and outcomes^5^. We demonstrated that both a PRS and a transcriptional risk score (TRS) independently predict COPD^6,7^. Although the PRS and TRS were both based on spirometry measures, the scores are not correlated^7^ and likely capture different aspects of lung pathobiology. Specifically, COPD genetic risk loci are enriched for aspects of lung development and have a greater effect in early COPD^6,8^; in contrast, the COPD TRS is associated with markers of inflammation and lung function decline and may reflect disease activity and propensity toward disease progression^7^. Thus, it may be possible to leverage the different features of the PRS and TRS to identify clinically and biologically distinct COPD subtypes.

Despite advances in omics-based risk prediction, important clinical translation questions remain. Omics risk scores are usually standardized for statistical analyses, leaving the question of how to use them to risk-stratify individuals^9^. Risk scores are also continuous measures, often normally distributed; the issue of attempting to identify subtypes along a continuum has been previously recognized^10^. Despite this limitation, there is a need to classify individuals to link omics-defined high-risk groups, which might benefit from specific therapies, with specific pathobiological processes and treatment decisions. COPD drug and drug repurposing candidates have high failure rates in clinical trials^11^, but it remains unknown if these therapies have failed because of patient selection, which currently does not utilize omics or other biomarkers.

The PRS was effective for predicting COPD severity and incident COPD, while the TRS was better at predicting FEV_1_ decline; combining both risk scores may identify subgroups at risk for multiple important COPD outcomes. Therefore, we hypothesized that our published PRS and TRS, both based on spirometry, could identify COPD subtypes (i.e., heterogeneity) within high-risk groups with clinical and biological differences in two cohorts of ever-smokers. We aimed to develop a novel approach for how omics risk scores can be applied to populations and leveraged for precision medicine. We used proteomics to obtain an additional biological view of omics-defined subgroups and performed in silico drug repurposing analyses to identify potential subgroup-specific drug repurposing candidates.

## METHODS

### Study populations

All study participants provided written informed consent and studies were approved by local Institutional Review Boards. Details regarding genotyping, RNA-sequencing, gene expression microarray, and proteomics data acquisition and processing are available in the Supplement.

#### COPDGene

We included Genetic Epidemiology of COPD (COPDGene) study (ClinicalTrials.gov Identifier: NCT00608764) participants with single nucleotide polymorphism (SNP) genotyping, RNA-sequencing, and SomaScan proteomic data to calculate the PRS, TRS, and performed differential protein expression analyses, respectively.

#### ECLIPSE

As previously described^7^, we included Evaluation of COPD Longitudinally to Identify Predictive Surrogate End-points (ECLIPSE) study (ClinicalTrials.gov Identifier: NCT00292552) participants with SNP genotyping data, whole blood microarray data, and at least two FEV_1_ measurements.

Additional details for both cohorts are in the supplement.

### Polygenic and transcriptional risk scores

The COPD PRS and TRS were both based on spirometry and previously described^6,7^; more details can be found in the supplement.

### Proteomic data

Blood proteomic data were measured using SomaScan v4.0, which uses aptamers (i.e. SOMAmers) to quantify 4,776 unique human proteins. Further details regarding SomaScan data and preparation can be found in the Supplement and here^12^.

### Statistical analysis

#### Overview of study design

To identify high-risk subgroups based on continuous scores, we used the same previously defined COPDGene training set^7^, and tested among PRS and TRS quantiles to maximize the number of associated differentially expressed proteins across the resulting subtype partitions (**Fig. 1**). We then determined the raw (non-standardized) score cut offs associated with the corresponding percentile values in the COPDGene training set. This approach facilitated classifying each participant in the COPDGene testing set and the external ECLIPSE validation set into an omics-defined subtype. We characterized the newly-defined subtypes using proteomic network and drug repurposing analyses, and applied multivariable linear regressions to test the association of subtypes with COPD-related outcomes.

**Figure 1:**
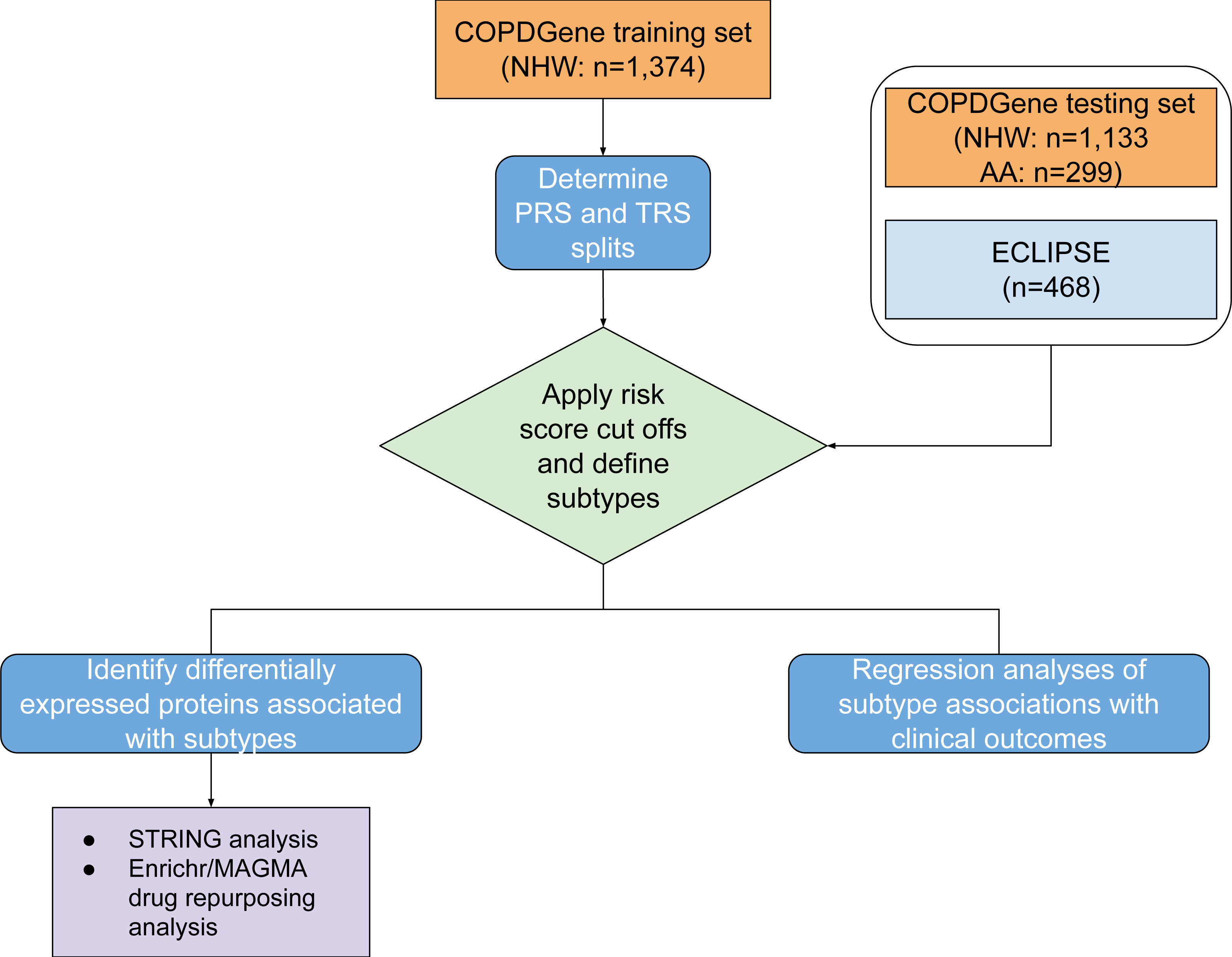
Schematic of study design. COPD = chronic obstructive pulmonary disease. COPDGene = Genetic Epidemiology of COPD study. ECLIPSE = Evaluation of COPD to Longitudinally Identify Predictive Surrogate Endpoints study. PRS = polygenic risk score. TRS = transcriptional risk score. STRING = Search Tool for the Retrieval of Interacting Genes/Proteins. MAGMA = Multi-marker Analysis of GenoMic Annotation.

#### Determining risk score divisions and identifying omics-defined subtypes

Omics-based risk scores are typically standardized prior to statistical analysis, and therefore, have a normal distribution and by design do not lend themselves to clustering analyses (**Figure S1**). Yet, for clinical application, patients need to be categorized into groups. First, to determine whether one or more clusters are optimal, we calculated the gap statistic based on the PRS and TRS using the clusGap function (cluster R package) with a maximum of 8 kmeans clusters and 500 bootstrap iterations. The gap statistic provides a measure of dispersion for each cluster and compares this dispersion metric to the expected dispersion under the null distribution^13^; thus, the difference (or “gap”) between the observed and expected within cluster dispersion is used to calculate the gap statistic and the maximum gap statistic over a range of cluster numbers indicates the optimal number of clusters.

As an alternative to clustering, individuals are commonly placed into omics risk-score quantiles, and participants in each quantile are compared to those in the lowest risk quantile^4,6,14^. To extend this approach to two separate omics risk scores is more complex, as fewer subdivisions lead to larger group sizes and more statistical power, but more subdivisions allow a more extreme comparison group. As genetic and transcriptomic data were used to define the subtypes, proteomics would provide a third “view” of the data. Thus, to determine the optimal quantiles we split the PRS and TRS into 2 to 3 quantiles (minimum of 4 and maximum of 9 groups) and tested to see what group divisions maximized the number of associated differentially expressed proteins using limma^15^, comparing each quantile category to the lowest quantile. To test the sensitivity of the groups to partitioning, we also examined clinical characteristics for each combination of these partitions. Benjamini-Hochberg^16^ false discovery rate (FDR)-adjusted p-values less than 0.05 were considered significant. The number of significantly differentially expressed proteins associated with each omics risk score category was summed.

#### Clinical comparisons of omics-defined subtypes

We compared clinical characteristics across omics-defined subtypes using the tableone R package. In COPDGene, transcriptomic and proteomic data were collected at the 5-year follow up visit (i.e., “Phase 2”), so we examined differences in anthropometry (including change in BMI per year (Kg/m^2^/year) from enrollment to the 5-year follow up visit), spirometry (including prospective FEV_1_ change (from 5-to 10-year follow up visits)), and CT measures of emphysema (quantitative emphysema on inspiratory CT scans (% LAA < -950 HU)^17^, 15th percentile of lung density histogram on inspiratory CT scans (Perc15)^18^) and of airway thickening (wall area percent (WA%)^17^ and square root of wall area of a hypothetical internal perimeter of 10 mm (Pi10)^19^) at the 5-year follow up visit. In ECLIPSE, we examined the same outcomes but longitudinal follow up was from the time of study enrollment to the 3-year follow up visit.

Further details regarding outcomes and regression model specifications are in the Supplementary Methods.

#### Biological characterization of omics-defined subtypes

We performed differential gene and protein expression analyses (accepting FDR-adjusted p-values <0.05), comparing high risk subtypes to the reference group, defined as the group with the lowest PRS and TRS quantiles. We mapped differentially expressed proteins to the human protein-protein interactome^20^ and performed Reactome^21^ pathway enrichment, STRING^24^ and Enrichr^22–24^ analyses; additional details are in the supplement.

## RESULTS

### Characteristics of study populations

We included 3,274 participants across two cohorts of individuals who smoked. The COPDGene training and testing sets are similar in demographic and spirometry characteristics (**Table 1**). Compared to COPDGene, ECLIPSE participants were more likely to be younger, male, to have a greater number of smoking pack-years, a lower FEV_1_ % predicted, and lower FEV_1_/FVC, and were less likely to be current smokers.

**Table 1:** Characteristics of study populations. COPD = chronic obstructive pulmonary disease. COPDGene = Genetic Epidemiology of COPD. ECLIPSE = Evaluation of COPD to Longitudinally Identify Predictive Surrogate Endpoints study. FEV_1_ = forced expiratory volume in 1 second. FEV_1_/FVC = FEV1/forced vital capacity. NHW = non-Hispanic white. AA=African American.

| *Characteristic* | *COPDGene training set* | *COPDGene testing set* |  | *ECLIPSE* |
| --- | --- | --- | --- | --- |
|  | *NHW* | *NHW* | *AA* | *NHW* |
| n | 1374 | 1133 | 299 | 468 |
| Age in years (mean (SD)) | 67.38 (8.25) | 68.13 (8.30) | 61.01 (6.95) | 64.43 (6.09) |
| Sex (No. % female) | 677 (49.3) | 560 (49.4) | 153 (51.2) | 156 (33.3) |
| Pack-years of smoking (mean (SD)) | 45.47 (24.61) | 45.47 (23.65) | 39.55 (20.74) | 49.33 (26.87) |
| Current smoking (No %) | 343 (25.0) | 289 (25.5) | 182 (60.9) | 70 (15.0) |
| FEV1 % predicted (mean (SD)) | 78.37 (24.21) | 77.85 (24.81) | 81.79 (23.54) | 44.22 (14.64) |
| FEV1/FVC (mean (SD)) | 0.67 (0.15) | 0.66 (0.15) | 0.71 (0.14) | 0.44 (0.11) |

### Defining polygenic and transcriptional risk score divisions

Participants plotted on the axes of PRS and TRS exist along a continuum (**Figure S1**), as is the case for spirometric measures of COPD severity (i.e., FEV_1_ and FEV_1_/FVC^10,25^). We calculated the gap statistic over a range of kmeans cluster numbers in the COPDGene training set, and consistent with visual inspection of **Figure S1**, we observed that one cluster yields the highest gap statistic, indicating that there are no clusters (**Figure S2**). As an alternative approach to clustering, we applied the common practice of dividing risk scores into quantiles, though the optimal quantiles balancing sufficiently high risk yet adequate sample size are not clear. Thus, we tested four combinations of partitioned omics risk scores, using protein expression differences (not used in the PRS and TRS) between groups. We observed that dichotomizing PRS and dividing TRS into tertiles yielded the greatest number of differentially expressed proteins (**Table S1**). We then applied these same quantiles to the COPDGene testing set and ECLIPSE participants (**Fig. 2**). To test the robustness of subgroups to specific partitions, we also examined 4 to 9 subdivisions, and noted stable clinical characteristics of the highest (“low PRS/high TRS” and “high PRS/high PRS”) and lowest risk (“low PRS/low TRS”) groups (**Table S2**).

**Figure 2.**
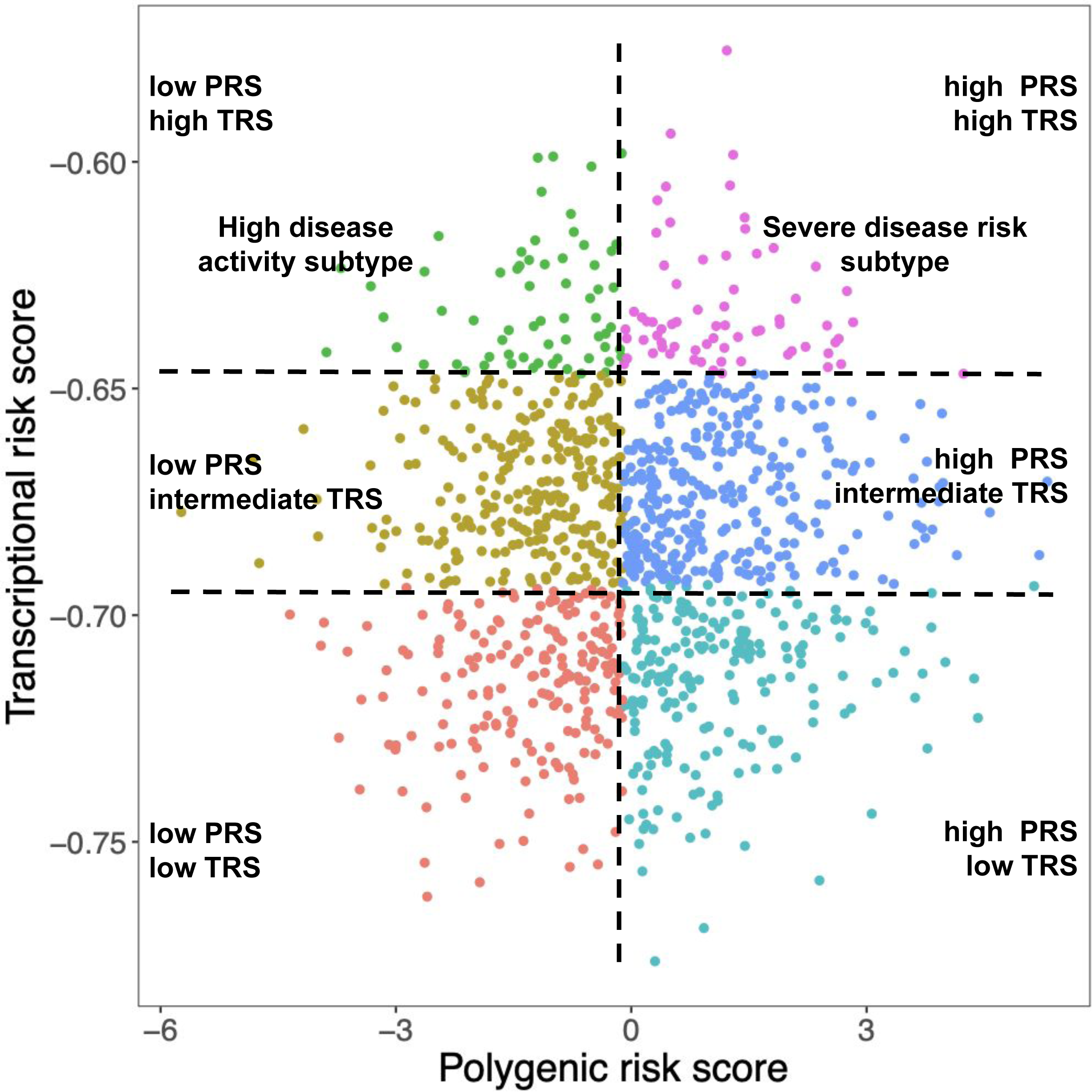
Omics-defined groups or subtypes overlaid on a plot of the polygenic risk score (PRS; *x-axis*) and transcriptional risk score (TRS; *y-axis*) in the COPDGene testing set.

### Polygenic and transcriptional risk scores identify “high disease activity” and “severe disease risk” subtypes

We observed, as expected, heterogeneity amongst the two high-risk (i.e., high TRS) groups (**Table 2**). We compared these high-risk subtypes to a reference group, which was defined as the lowest omics risk group (i.e., “low PRS/low TRS” subtype). Compared to the reference group, the two high-TRS risk groups (i.e., “low PRS/high TRS” and “high PRS/high TRS”) demonstrated decreased BMI, lower spirometry measures, more emphysema, and thicker airways across testing cohorts (**Table 2**). Both groups had similar mean adjusted prospective FEV_1_ decline in the COPDGene testing set compared to the reference group, but this finding was only consistent for the “low PRS/high TRS” group in ECLIPSE (-40 mL/year).

**Table 2:** Omics-defined subtypes in the COPDGene testing set and ECLIPSE. The was defined as the “Low PRS, Low TRS” group. BMI = body-mass index. LAA = low attenuation area. HU = Hounsfeld units. Perc15 = 15th percentile of lung density histogram on inspiratory CT scans. WA % = wall area percent. Pi10 = square root of wall area of a hypothetical internal perimeter of 10 mm. ACO = asthma-COPD overlap. See Table 1 legend for other abbreviations. NHW = non-Hispanic white. AA=African American.

| Characteristic | COPDGene NHW testing set |  |  | COPDGene AA testing set |  |  | ECLIPSE |  |  |
| --- | --- | --- | --- | --- | --- | --- | --- | --- | --- |
|  | Reference group<br>(Low PRS, Low TRS) | Low PRS,<br>High TRS | High PRS,<br>High TRS | Reference group<br>(Low PRS, Low TRS) | Low PRS,<br>High TRS | High PRS,<br>High TRS | Reference group (Low PRS, Low TRS) | Low PRS,<br>High TRS | High PRS,<br>High TRS |
| n | 196 | 65 | 68 | 61 | 16 | 11 | 57 | 63 | 56 |
| Age in years (mean (SD)) | 67.85 (8.52) | 67.16 (6.46) | 69.04 (8.11) | 61.26 (7.55) | 60.58 (7.24) | 65.95 (8.05) | 63.00 (5.10) | 65.43 (6.65) | 66.59 (5.95) |
| Sex (No % female) | 114 (58.2) | 19 (29.2) | 24 (35.3) | 36 (59.0) | 5 (31.2) | 8 (72.7) | 30 (52.6) | 10 (15.9) | 11 (19.6) |
| Pack-years of smoking (mean (SD)) | 36.51 (22.27) | 60.07 (26.49) | 57.28 (23.35) | 37.01 (20.85) | 43.42 (16.14) | 61.36 (38.87) | 45.49 (26.24) | 55.29 (35.25) | 54.29 (21.10) |
| Current smoking (No. %) | 20 (10.2) | 36 (55.4) | 36 (52.9) | 31 (50.8) | 15 (93.8) | 8 (72.7) | 6 (10.5) | 7 (11.1) | 12 (21.4) |
| BMI (Kg/m<sup>2</sup>) (mean (SD)) | 30.00 (6.84) | 26.51 (5.81) | 25.70 (5.58) | 30.61 (6.36) | 29.14 (8.52) | 25.26 (6.57) | 27.54 (5.06) | 26.70 (6.22) | 25.49 (4.76) |
| FEV1 % predicted (mean (SD)) | 90.15 (18.13) | 66.50 (27.07) | 60.55 (24.11) | 90.83 (17.41) | 80.61 (25.43) | 71.08 (10.67) | 50.25 (13.42) | 43.41 (15.65) | 37.44 (13.44) |
| FEV1/FVC (mean (SD)) | 0.75 (0.09) | 0.57 (0.15) | 0.53 (0.15) | 0.77 (0.10) | 0.73 (0.14) | 0.61 (0.12) | 45.84 (11.13) | 43.16 (12.79) | 39.71 (10.29) |
| % LAA < -950 HU (mean (SD)) | 2.78 (4.71) | 8.50 (10.39) | 11.00 (11.98) | 1.71 (4.00) | 3.51 (8.08) | 5.49 (7.30) | 16.01 (11.46) | 20.88 (13.50) | 22.44 (13.22) |
| Perc15 (mean (SD)) | -913.62 (20.43) | -927.23 (27.21) | -931.05 (31.55) | -895.53 (26.89) | -901.19 (29.94) | -913.62 (34.95) | -943.75 (50.13) | -960.00 (49.44) | -971.11 (45.84) |
| Pi10 (mean (SD)) | 2.01 (0.50) | 2.55 (0.65) | 2.60 (0.51) | 2.11 (0.48) | 2.56 (0.59) | 2.42 (0.44) | 4.38 (0.20) | 4.42 (0.23) | 4.43 (0.19) |
| WA % (mean (SD)) | 46.97 (8.11) | 54.46 (8.35) | 54.61 (7.41) | 47.48 (7.76) | 54.86 (10.52) | 51.62 (4.06) | 64.35 (2.79) | 65.05 (3.58) | 67.22 (3.44) |
| Change in FEV1 mL/year (prospective) (mean (SD)) | -32.11 (48.07) | -51.21 (74.41) | -66.45 (61.58) | -42.28 (36.59) | -87.86 (156.98) | -56.26 (76.62) | -32.21 (59.54) | -40.13 (75.31) | -15.22 (67.46) |

We then performed linear regression analyses on selected COPD-related outcomes. We compared anthropometric, spirometry, CT, and other COPD-related outcomes across COPDGene and ECLIPSE (**Table 3, Table S3**). Compared to the reference group, high-risk groups exhibited lower spirometry, more emphysema, and thicker airways (**Table 3**). While none of the adjusted FEV_1_ decline measures were statistically significant, the “low PRS, high TRS” means were consistent between COPDGene and ECLIPSE (-30ml/yr and -24ml/year, P = 0.15 and P = 0.11, respectively), despite that COPDGene participants had only two FEV_1_ measurements while ECLIPSE participants had up to six FEV_1_ measurements. Given the clinical relevance of accelerated FEV_1_ decline in the “low PRS/high TRS” group, we renamed this group the “high disease activity” subtype. While the “high PRS/high TRS” group did not have replicable FEV_1_ decline across cohorts, this group had the lowest lung function and most emphysema; therefore, we renamed this group the “severe disease risk” subtype. In COPDGene NHW participants only, the “high disease activity” subtype also exhibited a trend toward greater decline in BMI compared to the reference group (β = -0.154 [95% CI: -0.333 to 0.0253], p=0.094).

**Table 3.**
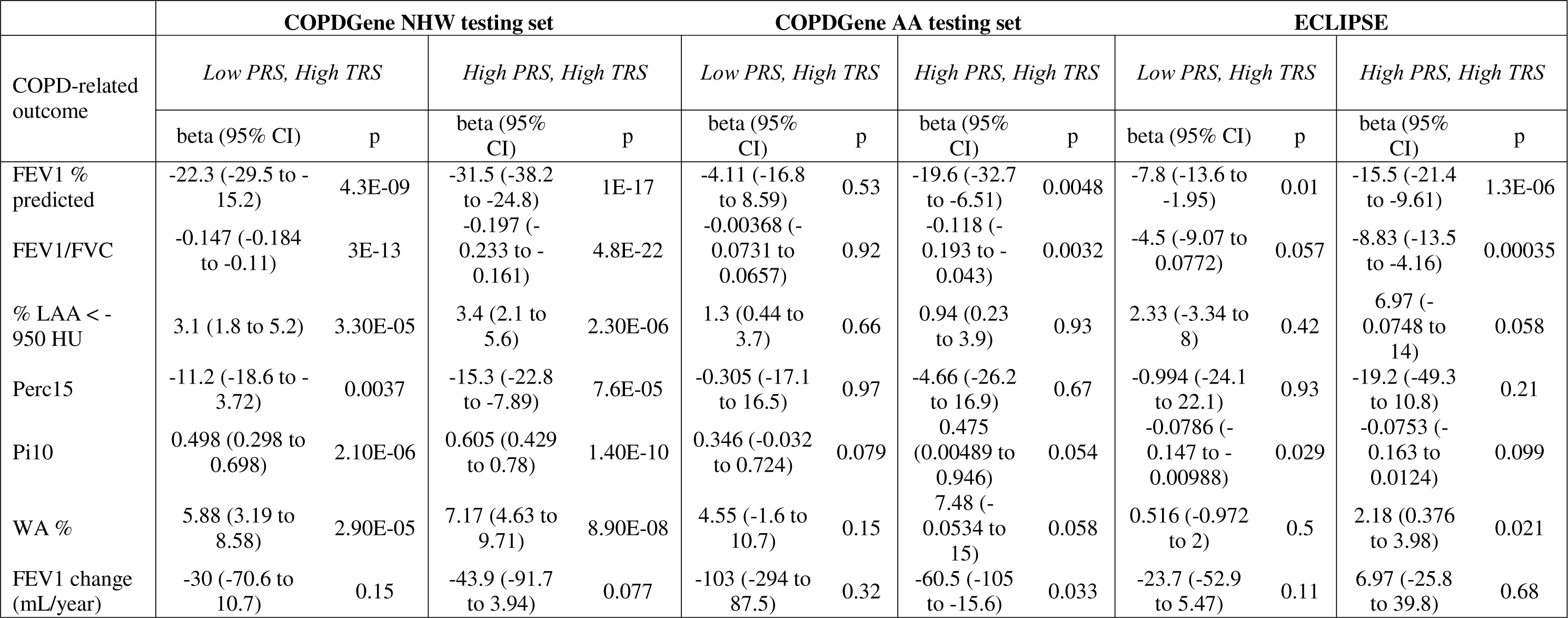
Multivariable linear regressions in the COPDGene testing sets and ECLIPSE. Models were adjusted or age, sex, current smoking status, pack-years of smoking, and principal components of genetic ancestry. Computed tomography imaging outcomes were additionally adjusted for CT scanner. Abbreviations are detailed in the Table 1 and 2 legends.

### Biological characterization and drug repurposing analyses of subtypes

Having identified clinical differences between the two high-risk groups, we sought to characterize biological differences between these subtypes. We performed differential gene and protein expression analyses between the “high disease activity” and “severe disease risk” subtypes and the reference group in the COPDGene NHW testing set (**Tables S4-S6**). The “high disease activity” subtype had 14 and the “severe disease risk” subtype had 2 differentially expressed proteins (**Table S6**). We did not observe differentially expressed genes or proteins when directly comparing high risk groups. We examined how the PRS affects differential gene expression associated with COPD case-control status as detailed in the supplement (Supplementary Results and **Table S7**)

We mapped differentially expressed proteins associated with each high-risk subtype in the COPDGene testing set to the human protein-protein interactome^20^ and used the mapped proteins as seed proteins to construct STRING PPI networks (**Figures 3 and 4**) with associated MCL clusters (**Table S8**) and perform pathway enrichment analyses (**Table S9**). To identify subtype-specific drug repurposing candidates, we used these same seed proteins to perform enrichment analyses on the MAGMA Drugs and Disease database^26^. Both subtypes demonstrated enrichment proteomic profiles suggesting potential treatment with ACE inhibitors, thyroid medications, carvedilol, bromocriptine, and lovastatin; the “high disease activity” subtype also had significant findings for 5-lipoxygenase inhibitors, fomepizole, and galantamine, while the “severe disease risk” (**Table S10**) subtype had significant findings for atypical antipsychotics.

**Figure 3.**
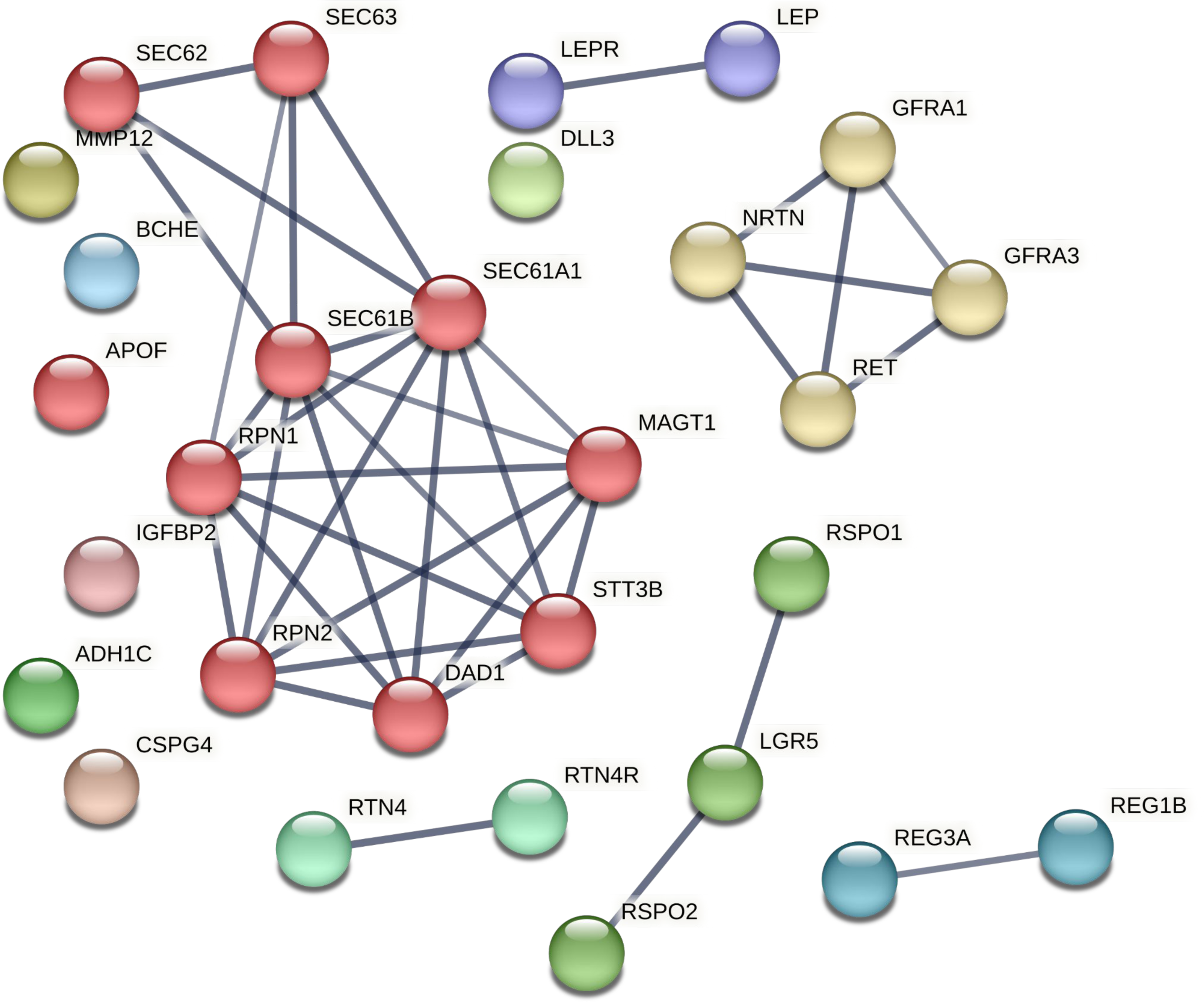
High disease activity (“low PRS, high TRS”) subtype STRING protein-protein interaction networks using differentially expressed proteins in Omics-defined groups (subtypes) in the COPDGene testing set as seed proteins, permitting up to 10 interactors in the first shell and 5 interactors in the second shell. Only high-confidence interactions were included and greater line thickness indicates greater confidence. Differentially expressed proteins were identified by comparing group assignments to the reference group. Colors represent MCL (Markov) clusters.

**Figure 4:**
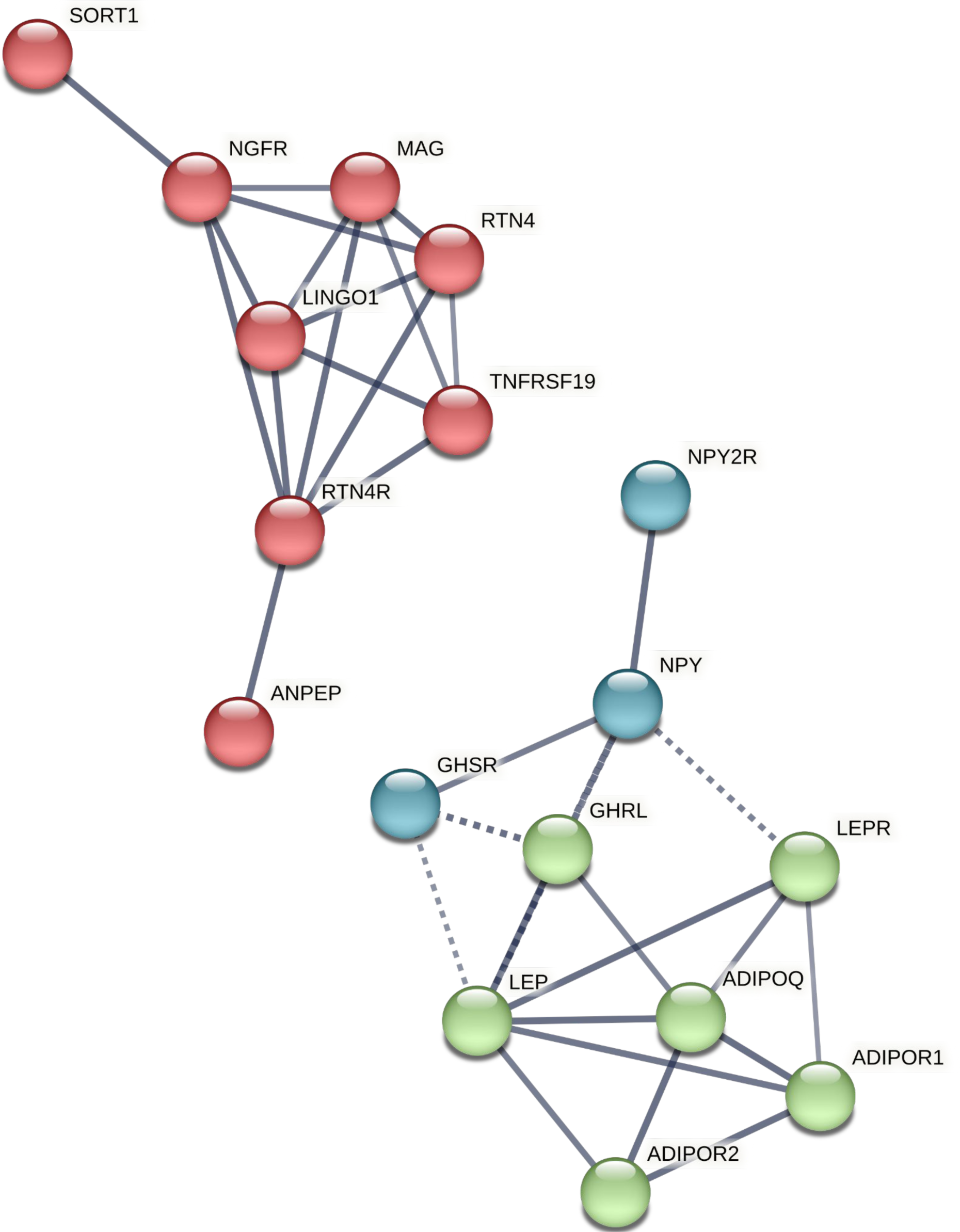
Severe disease risk (“high PRS, high TRS”) subtype STRING protein-protein interaction networks using differentially expressed proteins in Omics-defined groups (subtypes) in the COPDGene testing set as seed proteins, permitting up to 10 interactors in the first shell and 5 interactors in the second shell. Only high-confidence interactions were included and greater line thickness indicates greater confidence. Differentially expressed proteins were identified by comparing group assignments to the reference group. Colors represent MCL (Markov) clusters.

## DISCUSSION

In this study of 3,274 ever-smokers from two cohorts, we used blood-based polygenic (PRS) and transcriptional (TRS) risk scores to identify heterogeneity within high-risk individuals, defining “high disease activity” and “severe disease risk” COPD subtypes. Compared to a reference group, both subtypes had lower mean BMI values and alterations in metabolic, growth, and immune signaling processes. “High disease activity” participants exhibited prospective FEV_1_ decline across both replication cohorts, albeit with a non-significant (though directionally consistent) association in multivariable models adjusted for baseline FEV_1_. “Severe disease risk” participants had low spirometry measures with high quantitative emphysema and thick airways. We identified biological processes and drug repurposing candidates associated with each subtype, including therapies previously tested in COPD clinical trials. Our study demonstrates how omics risk scores can identify COPD subtypes with associated clinical and biological characteristics that can be leveraged for therapeutic interventions.

Linking omics-defined high-risk groups to specific pathobiology is an active area of research that we now extend to lung disease. In schizophrenia, PRS-defined high risk groups were biologically characterized using weighted gene co-expression network analyses^27^. Other approaches have incorporated gene expression into PRSs to improve prediction and imply biological mechanisms^28–30^. Here, we used genetic and transcriptomic data to define subtypes, and then leveraged proteomic differences between groups to understand subtype biology. Importantly, our results suggest that different omics risk scores are not interchangeable, i.e., a higher omics risk score will not always have the same association with specific outcomes. Individuals with the highest transcriptomic quantile exhibited different clinical and biological features depending on their underlying polygenic risk. We also demonstrate that individuals may exist along a continuum of COPD risk – i.e., there are no clusters – yet omics-defined subgroups may have important clinical and biological differences. While we did not observe clusters, we observed that omics-defined subgroups were robust to varying PRS and TRS subdivisions, with the two high-risk groups (“low PRS/high TRS” and “high PRS/high TRS”) demonstrating stable clinical characteristics across risk score subdivisions. Thus, genetics and transcriptomics may provide alternative yet complementary views of lung function biology.

Of direct clinical relevance, the association of lower spirometry, greater emphysema and observed FEV_1_ decline across two cohorts suggests that the “high disease activity” subtype is a targetable trait, the risk of which, might be modified by approved medications (5-lipoxygenase inhibitors, angiotensin-converting enzyme (ACE) inhibitors, fomepizole, galantamine). Fomepizole has not previously been implicated as a COPD drug repurposing candidate to our knowledge. Galantamine is known to cause bronchospasm, and enrichment for proteins targeted by galantamine suggests this drug is most likely to cause harm in this subtype of patients. Although a randomized trial of the 5-lipoxygenase inhibitor, Zileuton, did not reduce length of stay or treatment failure in patients hospitalized for COPD exacerbations, it was likely underpowered^31^, and did not examine longer term outcomes. ACE inhibitors and angiotensin receptor blockers (ARB) have been identified as COPD drug repurposing candidates. A recent clinical trial showed failure of losartan to decrease emphysema progression^32^. Our drug repurposing analyses implicated utility of captopril – not all ACE inhibitors and ARBs - in the “high disease activity” but not the “severe disease risk” subtype. Conversely, our analysis suggests potential benefit of atypical antipsychotics in the “severe disease risk” subtype. The coincidence of schizophrenia and COPD is largely attributed to smoking, though a phenome-wide association and polygenic risk analysis suggests that schizophrenia and obstructive lung disease may have shared genetic mechanisms^33^. While we adjusted for self-reported cigarette smoking status, these measures are imperfect, and thus we cannot address whether shared mechanisms are due to smoking behavior. A broader implication of the drug repurposing analyses that merits validation is that previous failure of drugs in clinical trials was due to heterogeneity in patient selection that could be overcome by omics-based subtyping.

The cachectic COPD patient, who may be prone to more exacerbations, is a well-described clinical phenotype ^34–37^, and we observed that the “high disease activity” and “severe disease risk” subtypes have lower mean BMI than other subtypes. Although pulmonary cachexia has been proposed to occur only in severe COPD^38^, we demonstrate additional heterogeneity within lower BMI ever-smokers and identify a “high disease activity” subtype with less spirometric severity and distinct biology. The “high disease activity” group may also overlap with a previously identified comorbidity cachexia subgroup^39^, and in the current analysis, we identified molecular profiles that might guide therapy. Relevant to body composition, the adipocyte product leptin exhibited altered expression in both subtypes, which has several potential, not mutually exclusive interpretations. Leptin acts both as a hormone negatively regulating hunger and adipocyte fat storage^40^ and as a proinflammatory cytokine essential for host defense^41,42^.

The role of leptin in regulating hunger suggests it could be relevant to the “severe disease risk” subtype, which had the largest effect size in leptin expression (-0.71 log-fold change); the observed decrease in leptin could be a compensatory response to or share a causal relationship with pulmonary cachexia. In addition, the lower leptin levels observed in malnourished populations are associated with dysfunctional cell-mediated immunity and increased susceptibility to infections^41,43^. In COPD, elevated leptin concentrations have been reported in the plasma and airways in some^44–48^, but not all studies^49^, and has been identified as a potential biomarker of emphysema progression^50,51^. Our results suggest that “high disease activity” or “severe disease risk” subtype individuals might exhibit humoral and cytokine profiles similar to those seen in malnourished individuals with increased susceptibility to infections. It remains unclear whether the observed peripheral blood proteomic alterations are merely a consequence of disease activity, or a causal component of a positive feedback loop involved in driving disease progression. The biomarkers used in this study are blood-based, making it difficult to identify the relevant mechanisms in lung tissue. However, blood-based biomarkers are practical in a clinical setting and follow up studies linking changes in these biomarkers to pathophysiologic mechanism in lung tissue can help to bridge the gap between prediction and precision therapeutics.

This study leveraged multiple omics data in the form of validated, replicated, risk scores, to identify COPD subtypes in two cohorts of ever-smokers. One major challenge of this analysis was determining how to subset subjects from a standardized, continuous distribution. To overcome this limitation, we varied the number of quantiles and assessed which combination of divisions yielded the greatest number of differentially expressed proteins, then identified corresponding raw score cut-off values that allowed each participant in the COPDGene and ECLIPSE testing sets to be categorized into an omics-defined subtype. We acknowledge that there are other reasonable approaches for applying omics risk scores to identify COPD subtypes.

Individuals near the cut-off value for PRS might exist on a continuum between “high disease activity” and “severe disease risk” subtypes, and these participants might be able to transition between subtypes; regardless, defining thresholds and categories are an important step toward clinical translation. We observed similar results in COPDGene AA participants, including the “high disease activity” subtype association with FEV_1_ decline in multivariable analyses; however, additional work to optimize multi-ancestry genetic and omics-based prediction is crucial to prevent omics technologies from becoming a vehicle for worsening existing healthcare disparities^52^. We based our drug repurposing candidates on enrichment analyses of proteomic profiles, but directionality of biological processes was not accounted for in enrichment analyses; additional mechanistic and clinical trial validations are needed. Finally, we identified two high-risk subtypes using this approach, but these do not explain all of COPD heterogeneity, and there are almost certainly other important subtypes.

In conclusion, polygenic and transcriptional risk scores, both based on spirometry, identified “high disease activity” and “severe disease risk” subtypes with distinct clinical and biological characteristics. Proteomic and drug repurposing analysis identified subtype-specific enrichment for therapies, some of which were previously hypothesized in COPD.

## Data Availability

All data are available through the database of Genotypes and Phenotypes (dbGaP)

## Author contributions

Study Design: Matthew Moll, Julian Hecker, Michael H. Cho, Brian D. Hobbs.

Acquisition, analysis, or interpretation of the data: Matthew Moll, Julian Hecker, Edwin K. Silverman, Auyon Ghosh, Don D. Sin, Peter J. Castaldi, Katherine Pratte, Russell Bowler, Brian D. Hobbs, Michael H. Cho.

Critical revision of the manuscript for important intellectual content: All authors

Statistical analysis: Matthew Moll, Julian Hecker, John Platig, Kimberly Glass, Brian D. Hobbs, Michael H. Cho.

Obtained funding: Edwin K. Silverman, Michael H. Cho, Matthew Moll

