## Supplementary appendix for "Polygenic and transcriptional risk scores identify chronic obstructive pulmonary disease subtypes"

### Funding and Acknowledgements

**COPDGene Phase 3 Grant Support and Disclaimer**

The project described was supported by NHLBI grants U01 HL089897 and U01 HL089856 and by NIH contract 75N92023D00011. The content is solely the responsibility of the authors and does not necessarily represent the official views of the National Heart, Lung, and Blood Institute or the National Institutes of Health.

**COPD Foundation Funding**

COPDGene is also supported by the COPD Foundation through contributions made to an Industry Advisory Board that has included AstraZeneca, Bayer Pharmaceuticals, Boehringer- Ingelheim, Genentech, GlaxoSmithKline, Novartis, Pfizer, and Sunovion.

**COPDGene® Investigators – Core Units**

*Administrative Center*: James D. Crapo, MD (PI); Edwin K. Silverman, MD, PhD (PI); Barry J. Make, MD; Elizabeth A. Regan, MD, PhD

*Genetic Analysis Center*: Terri H. Beaty, PhD; Peter J. Castaldi, MD, MSc; Michael H. Cho, MD, MPH; Dawn L. DeMeo, MD, MPH; Adel El Boueiz, MD, MMSc; Marilyn G. Foreman, MD, MS; Auyon Ghosh, MD; Lystra P. Hayden, MD, MMSc; Craig P. Hersh, MD, MPH; Jacqueline Hetmanski, MS; Brian D. Hobbs, MD, MMSc; John E. Hokanson, MPH, PhD; Wonji Kim, PhD; Nan Laird, PhD; Christoph Lange, PhD; Sharon M. Lutz, PhD; Merry-Lynn McDonald, PhD; Dmitry Prokopenko, PhD; Matthew Moll, MD, MPH; Jarrett Morrow, PhD; Dandi Qiao, PhD; Elizabeth A. Regan, MD, PhD; Aabida Saferali, PhD; Phuwanat Sakornsakolpat, MD; Edwin K. Silverman, MD, PhD; Emily S. Wan, MD; Jeong Yun, MD, MPH

*Imaging Center*: Juan Pablo Centeno; Jean-Paul Charbonnier, PhD; Harvey O. Coxson, PhD; Craig J. Galban, PhD; MeiLan K. Han, MD, MS; Eric A. Hoffman, Stephen Humphries, PhD; Francine L. Jacobson, MD, MPH; Philip F. Judy, PhD; Ella A. Kazerooni, MD; Alex Kluiber; David A. Lynch, MB; Pietro Nardelli, PhD; John D. Newell, Jr., MD; Aleena Notary; Andrea Oh, MD; Elizabeth A. Regan, MD, PhD; James C. Ross, PhD; Raul San Jose Estepar, PhD; Joyce Schroeder, MD; Jered Sieren; Berend C. Stoel, PhD; Juerg Tschirren, PhD; Edwin Van Beek, MD, PhD; Bram van Ginneken, PhD; Eva van Rikxoort, PhD; Gonzalo Vegas Sanchez- Ferrero, PhD; Lucas Veitel; George R. Washko, MD; Carla G. Wilson, MS;

*PFT QA Center, Salt Lake City, UT*: Robert Jensen, PhD

*Data Coordinating Center and Biostatistics*, *National Jewish Health, Denver, CO*: Douglas Everett, PhD; Jim Crooks, PhD; Katherine Pratte, PhD; Matt Strand, PhD; Carla G. Wilson, MS

*Epidemiology Core*, *University of Colorado Anschutz Medical Campus, Aurora, CO*: John E. Hokanson, MPH, PhD; Erin Austin, PhD; Gregory Kinney, MPH, PhD; Sharon M. Lutz, PhD; Kendra A. Young, PhD

*Mortality Adjudication Core:* Surya P. Bhatt, MD; Jessica Bon, MD; Alejandro A. Diaz, MD, MPH; MeiLan K. Han, MD, MS; Barry Make, MD; Susan Murray, ScD; Elizabeth Regan, MD; Xavier Soler, MD; Carla G. Wilson, MS

*Biomarker Core*: Russell P. Bowler, MD, PhD; Katerina Kechris, PhD; Farnoush Banaei- Kashani, PhD

**COPDGene® Investigators – Clinical Centers**

*Ann Arbor VA:* Jeffrey L. Curtis, MD; Perry G. Pernicano, MD

*Baylor College of Medicine, Houston, TX*: Nicola Hanania, MD, MS; Mustafa Atik, MD; Aladin Boriek, PhD; Kalpatha Guntupalli, MD; Elizabeth Guy, MD; Amit Parulekar, MD;

*Brigham and Women’s Hospital, Boston, MA*: Dawn L. DeMeo, MD, MPH; Craig Hersh, MD, MPH; Francine L. Jacobson, MD, MPH; George Washko, MD

*Columbia University, New York, NY*: R. Graham Barr, MD, DrPH; John Austin, MD; Belinda D’Souza, MD; Byron Thomashow, MD

*Duke University Medical Center, Durham, NC*: Neil MacIntyre, Jr., MD; H. Page McAdams, MD; Lacey Washington, MD

*HealthPartners Research Institute, Minneapolis, MN*: Charlene McEvoy, MD, MPH; Joseph Tashjian, MD

*Johns Hopkins University, Baltimore, MD*: Robert Wise, MD; Robert Brown, MD; Nadia N. Hansel, MD, MPH; Karen Horton, MD; Allison Lambert, MD, MHS; Nirupama Putcha, MD, MHS

*Lundquist Institute for Biomedical Innovation at Harbor UCLA Medical Center, Torrance, CA*: Richard Casaburi, PhD, MD; Alessandra Adami, PhD; Matthew Budoff, MD; Hans Fischer, MD; Janos Porszasz, MD, PhD; Harry Rossiter, PhD; William Stringer, MD

*Michael E. DeBakey VAMC, Houston*, *TX*: Amir Sharafkhaneh, MD, PhD; Charlie Lan, DO *Minneapolis VA:* Christine Wendt, MD; Brian Bell, MD; Ken M. Kunisaki, MD, MS

*Morehouse School of Medicine, Atlanta, GA*: Eric L. Flenaugh, MD; Hirut Gebrekristos, PhD; Mario Ponce, MD; Silanath Terpenning, MD; Gloria Westney, MD, MS

*National Jewish Health, Denver, CO*: Russell Bowler, MD, PhD; David A. Lynch, MB *Reliant Medical Group, Worcester, MA*: Richard Rosiello, MD; David Pace, MD

*Temple University, Philadelphia, PA:* Gerard Criner, MD; David Ciccolella, MD; Francis Cordova, MD; Chandra Dass, MD; Gilbert D’Alonzo, DO; Parag Desai, MD; Michael Jacobs, PharmD; Steven Kelsen, MD, PhD; Victor Kim, MD; A. James Mamary, MD; Nathaniel Marchetti, DO; Aditi Satti, MD; Kartik Shenoy, MD; Robert M. Steiner, MD; Alex Swift, MD; Irene Swift, MD; Maria Elena Vega-Sanchez, MD

*University of Alabama, Birmingham, AL:* Mark Dransfield, MD; William Bailey, MD; Surya P. Bhatt, MD; Anand Iyer, MD; Hrudaya Nath, MD; J. Michael Wells, MD

*University of California, San Diego, CA*: Douglas Conrad, MD; Xavier Soler, MD, PhD; Andrew Yen, MD

*University of Iowa, Iowa City, IA*: Alejandro P. Comellas, MD; Karin F. Hoth, PhD; John Newell, Jr., MD; Brad Thompson, MD

*University of Michigan, Ann Arbor, MI:* MeiLan K. Han, MD MS; Ella Kazerooni, MD MS; Wassim Labaki, MD MS; Craig Galban, PhD; Dharshan Vummidi, MD

*University of Minnesota, Minneapolis, MN*: Joanne Billings, MD; Abbie Begnaud, MD; Tadashi Allen, MD

*University of Pittsburgh, Pittsburgh, PA*: Frank Sciurba, MD; Jessica Bon, MD; Divay Chandra, MD, MSc; Joel Weissfeld, MD, MPH

*University of Texas Health, San Antonio, San Antonio, TX*: Antonio Anzueto, MD; Sandra Adams, MD; Diego Maselli-Caceres, MD; Mario E. Ruiz, MD; Harjinder Singh

The **ECLIPSE** study (NCT00292552; GSK code SCO104960) was funded by GlaxoSmithKline.

### Supplementary Methods

#### Additional cohort details

##### COPDGene

Briefly, the COPDGene study recruited n=10,198 non-Hispanic white (NHW) and African American (AA) individuals aged 45-80 years with ≥10 pack-years of smoking history^1^. COPDGene began as a cross-sectional case-control study that was extended into a longitudinal study including 5- and 10-year follow up visits. Anthropometric, spirometry, and computed tomography (CT) imaging measures were performed at each visit. We obtained genotype data at baseline, and RNA-sequencing and SomaScan proteomic data at the 5-year follow up visit.

Single nucleotide polymorphism (SNP) genotyping was performed using the Illumina (San Diego, CA) HumanOmniExpress array. Genotyping at the Z and S alleles was performed and participants with severe alpha-1 antitrypsin deficiency were excluded. Imputation was performed using the Michigan Imputation Server to the Haplotype Reference Consortium^1^ and 1000 Genomes Phase I v3 Cosmopolitan reference panels, for non-Hispanic whites and African Americans, respectively. Variants with an r^2^ value of ≤ 0.3 were removed.

##### ECLIPSE

The ECLIPSE study recruited n=2,140 individuals with COPD aged 40-75 years and ≥10 pack-years of smoking history^2^. Baseline anthropometric, spirometry, and CT imaging measures were collected. Blood samples were also collected at study enrollment and, for a subset of samples, both genotype and gene expression microarray data are available. If individuals met the spirometry criteria for Global Initiative for Chronic Obstructive Lung Disease (GOLD)^3^ stage 2-4 COPD at enrollment, they returned every six months for three years for repeat spirometry.

In ECLIPSE, SNP genotyping was performed using the Illumina HumanHap 550 V3 (Illumina, San Diego, CA) array. Subjects and markers with call rates < 95% were excluded. Imputation was performed using the Michigan Imputation Server and Haplotype Reference Consortium^1^ reference panel.

#### Cohort expression data

##### COPDGene: RNA sequencing data

At the 5-year follow up of the COPDGene study, whole blood was obtained and stored in PAXgene Blood RNA tubes. Collection and processing of RNA-sequencing data was previously described^2^. Briefly, total RNA was extracted with the Qiagen PreAnalytiX PAXgene Blood miRNA Kit (Qiagen, Valencia, CA). After undergoing quality assurance, samples were globin reduced and cDNA library preparation was performed; 75 bp reads with a mean of 20 million reads per sample were generated using an Illumina HiSeq 2500. Count data were filtered to include transcripts with > 1 count per million (CPM) in 99% of samples, and were subsequently normalized by log-CPM transformation using the edgeR R package^3^. Counts were adjusted for library depth, and batch effects were removed using the limma removeBatchEffects function^4^.

##### ECLIPSE: Microarray data

ECLIPSE participants had blood samples collected at the time of study enrollment, and total RNA was extracted using PAXgene Blood miRNA kits and hybridized to the Affymetrix Human Gene 1.1 ST array. If transcripts were represented by multiple probes, we chose the probe with the greatest interquartile range. Batch effects were removed using the limma removeBatchEffects function^4^. Please refer to our prior publication^2^ for further details regarding preparation and processing of RNA data.

Prior to analyses, we limited transcripts to those present in both data sets based on HGNC symbols. For ECLIPSE microarray data, some gene transcripts were represented by multiple probes. In these cases, we chose the probe with the greatest interquartile range. We also scaled the RNAseq count and microarray gene expression data to have a mean of 0 and standard deviation of 1.

##### COPDGene: SomaScan Proteomic data

Using the SomaScan 5K platform, we performed plate hybridization, median signal normalization, and plate scaling and calibration of SOMAmers to control for variability across array signals, inter-run variability, inter-assay variation between analytes and batch differences between plates. Further details regarding preparation of SomaScan data has been previously published.

#### Additional statistical analyses

##### Polygenic risk score

We previously published a PRS^4^ using GWASs of FEV_1_ and FEV_1_/FVC performed in approximately 500,000 individuals from the UK Biobank and SpiroMeta consortium^5^. We calculated PRSs for FEV_1_ and FEV_1_/FVC separately using lassosum^6^, a penalized regression approach that minimizes collinearity, provides feature selection, and accounts for linkage disequilibrium. We summed the FEV_1_ and FEV_1_/FVC scores into a composite risk score, as previously performed^4^. COPDGene and ECLIPSE were not used to develop the PRS and represent external datasets.

To maximize our ability to separate subjects based on the genetic risk score and to minimize potential confounding by genetic ancestry, we analyzed NHW and AA participants separately in this analysis and residualized by regressing out principal components of genetic ancestry from the PRS before use.

##### Transcriptional risk score

We previously published a TRS^7^ in a training sample of COPDGene using least absolute shrinkage and selection operator (LASSO) penalized regression^8^ in 1,374 individuals from the COPDGene study and tested its performance in a held-out sample of 674 individuals. We have since obtained RNA-sequencing data (TOPMed Freeze 4) in an additional 459 NHW and 143 AA participants and have added these participants to the COPDGene testing set. We ensured that none of the samples used in the training of the TRS were included in our COPDGene testing set.

##### Clinical comparisons of omics-defined subtypes

We defined FEV_1_ change in COPDGene as the 10-year follow-up measure minus the 5-year follow-up measure, divided by the time in years, and in ECLIPSE, by taking the slope of the best fit line for FEV_1_ versus time, as previously described^7,9^. In COPDGene, we additionally examined differences in early-onset COPD (GOLD 2-4 spirometry grades before 55 years of age^10^) and absolute white blood cell differential counts. We tested the interaction of the PRS and TRS for each outcome by including both main effects and the cross-product term in a regression model (interaction term = PRS X TRS).

##### Regression model specifications

We performed multivariable linear regressions in the COPDGene testing set comparing each subtype to the reference group. Outcomes included FEV_1_ % predicted, FEV_1_/FVC, % LAA < -950 HU, Perc15, Pi10, and WA% . We adjusted regression models for age, sex, current smoking status, pack-years of smoking history, principal components of genetic ancestry, and CT scanner (for imaging outcomes only). We selected outcomes based on input from clinicians and data availability (less than 20% missingness). Adjustment variables were chosen based on clinician input and these measures were available for all participants. We additionally performed interaction analyses by including the PRS, TRS, and a cross-product term (PRS*TRS) within a single regression model.

Biological characterization of omics-defined subtypes

##### Differential expression analysis

We performed differential protein expression analysis to identify proteins associated with the PRS. As described in the main text, we used limma and considered FDR-adjusted p-values below 0.05 to be significant. We also compared differential protein expression between the high-risk groups. To understand how the PRS modifies gene expression profiles, we also examined differentially expressed genes associated with COPD case-control status (GOLD 2-4 versus normal spirometry) adjusting for age, sex, smoking status, pack-years of smoking. We then repeated this analysis adjusting for the PRS and principal components of genetic ancestry. We chose adjustment variables based on clinician input and the fact that these measures were available for all participants.

##### STRING, pathway enrichment, and drug repurposing analyses

We used STRING ([www.string-db.org](http://www.string-db.org)) to query the human protein-protein interactome and to construct a network including up to 10 interactors in the first shell (i.e., proteins directly interacting with seed proteins) and 5 interactors in the second shell (i.e., proteins directly interacting with 1^st^ shell proteins) per seed protein^11^; only high confidence interactions (≥ 0.7) were included.

We also performed pathway enrichment and MCL (Markov clustering algorithm, inflation parameter 3) clustering analyses^12^ on these protein-protein interaction (PPI) networks. We input the differentially expressed proteins into Enrichr (maayanlab.cloud/Enrichr) to query the Multi-marker Analysis of GenoMic Annotation (MAGMA) drugs and diseases database^13^, and used the Enrichr Appyter to identify potential drug repurposing candidates for individuals belonging to omics-defined subtypes. After these analyses, we renamed the subtype groups based on associated clinical outcomes and biological processes.

### Supplementary Results

#### *Defining polygenic and transcriptional risk score divisions*

Thus, an individual’s PRS category was defined by the relation of their score to the raw score value corresponding to the 50^th^ percentile in the COPDGene training set (a non-standardized PRS value of -0.11, see Supplement). TRS values were analogously categorized based on the 33^rd^ and 66^th^ percentiles (“low TRS”: non-standardized TRS < -0.69; “intermediate TRS”: non-standardized TRS < -0.65).

#### *Polygenic and transcriptional risk scores identify ‘high disease activity’ and ‘severe disease’ subtypes*

In interaction analyses, the PRS*TRS cross-product term was not significantly associated with any of the tested outcomes (all p > 0.05).

#### *Biological characterization and drug repurposing analyses of subtypes*

COPD case-control status was associated with 6,826 genes, adjusting for clinical risk factors; adding the PRS as a covariate altered the gene expression profile to include 6,523 genes, 6,168 of which were present in both analyses while 355 genes were unique to the analysis adjusted for the PRS. The top 10 differentially expressed genes (based on log-fold change) for the clinical model and clinical model with PRS were similar and involved genes involved in response to infections and RNA regulatory elements (**Table S7**).

*Biological characterization and drug repurposing analyses of subtypes*

Leptin signaling was negatively associated (negative direction of effect) with both high-risk subtypes (**Table S6**). The ‘high disease activity’ subtype was associated with enrichment for maturation of spike protein, SRP-dependent cotranslational protein targeting to membrane, and regulation of frizzled (FZD) by ubiquitination pathways (**Table S9**). The ‘severe disease risk’ subtype was enriched for proteins in AMP-activated protein kinase (AMPK) inhibition of chREBP, peptide hormone synthesis, and Ghrelin signaling pathways (**Table S9**).

#### Supplementary Figures

Figure S1. Participants plotted on the axes of the polygenic (PRS) (x-axis) and transcriptional (TRS) (y-axis) risk scores. Both risk scores are standardized to mean of 0 and standard deviation of 1.

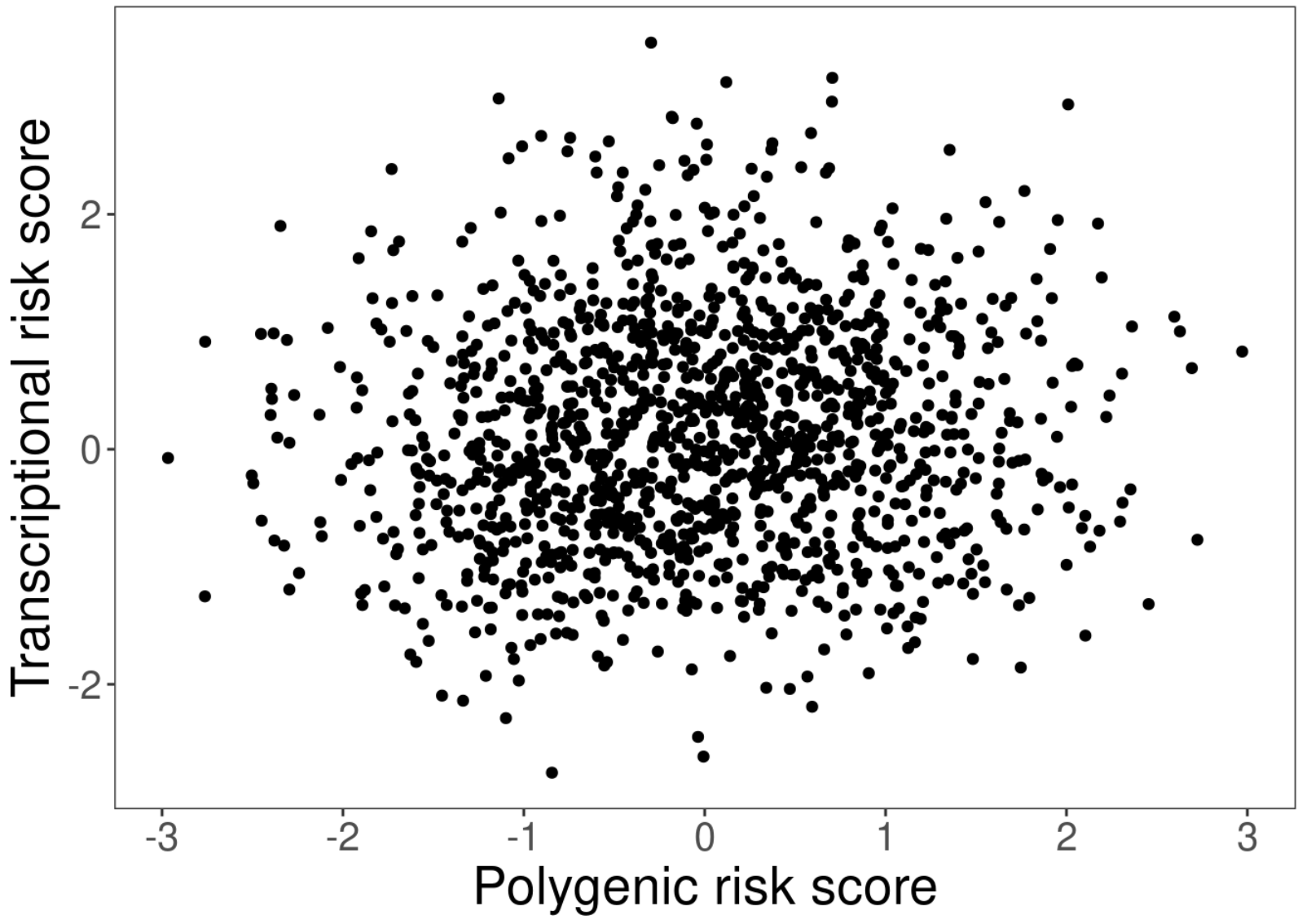

Figure S2: The gap statistic^14^ for kmeans clusters calculated based on the polygenic and transcriptional risk scores in the COPDGene training set with 100 bootstrap iterations calculated for 1 to 8 clusters. A higher gap statistic indicates a more optimal cluster number.

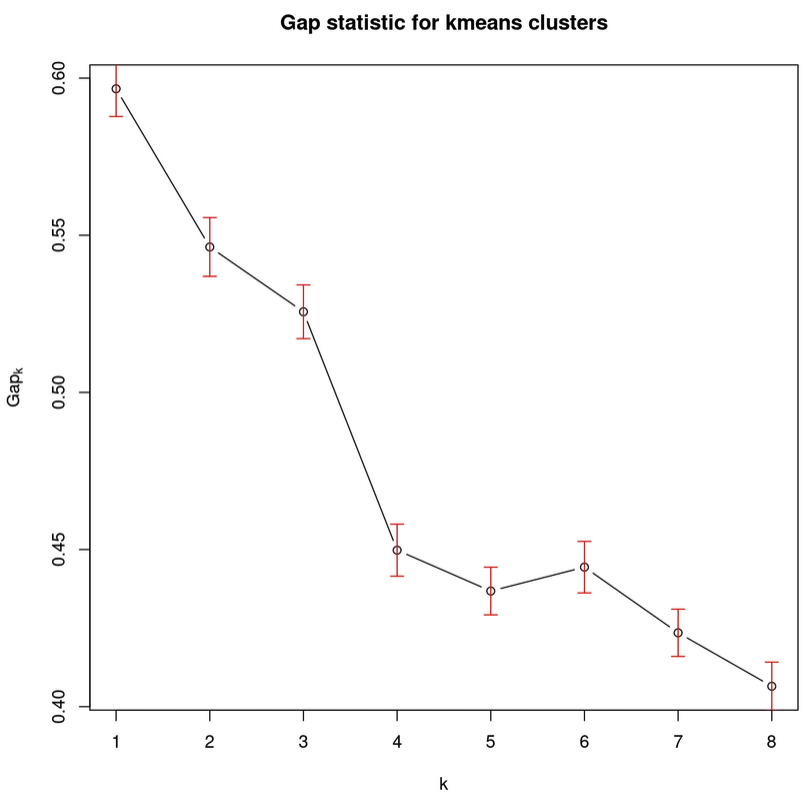

#### Supplementary Tables

See online supplementary file: supplementary_tables.xlsx

### Supplementary Discussion

We observed that the ‘high disease activity’ subtype, characterized by rapid FEV_1_ decline (-30 mL/year in COPDGene), had alterations in responses to infections (i.e., spike protein maturation) and regulation of frizzled (FZD*)* Wnt receptors by ubiquitination pathways. While no direct link between SARS-CoV-2 and FEV_1_ decline has been reported, the follow up time is limited for such studies. Downregulation of FZD and decreased Wnt/β-catenin signaling has been observed in COPD patients’ alveolar cells^15^. In a GWAS follow up study of a *FAM13A* COPD risk variant, Wnt/β-catenin signaling was downregulated and associated with emphysema in mice^16^. Taken together, this subtype appears to exhibit alterations in protein signaling that reflect responses to infections and signaling pathways implicated in emphysema.

By contrast, different pathways (AMPK inhibition of chREBP, peptide hormone synthesis, and Ghrelin signaling pathways) were associated with the ‘severe disease risk’ subtype, which had more emphysema and airway pathology, and lower spirometry measures, yet no significant difference from the reference group in FEV_1_ decline. Ghrelin is secreted by the stomach and stimulates appetite and weight gain and has been observed to be increased only in underweight COPD patients, presumably as a compensatory mechanism^17^. Metformin-mediated AMPK activation reduced elastase-induced emphysema, inflammatory responses, and markers of cellular senescence in mice^18^. Interestingly, drug repurposing analyses did not implicate metformin as a subtype-specific therapy, suggesting that the enrichment for proteins associated with metformin were not overrepresented enough to pass multiple comparison testing. Given that this subtype has a lower mean BMI compared to other groups and has alterations in other metabolic signaling pathways, it is possible that this subtype could benefit from metformin use, though additional investigation is certainly needed.

In addition, using raw score cut-offs to categorize individuals may complicate transferability to general populations. However, concern about homogeneity of our study populations is reduced because the PRS was derived from a general population cohort from which COPDGene and ECLIPSE differ considerably in smoking severity and disease severity; COPDGene and ECLIPSE also differ from each other with respect to disease severity and method of RNA assay (RNA-seq and microarray, respectively). Another limitation is uncertainty on the optimal time to apply such a classification schema, as genetics can be measured at birth, but transcriptomics change over a lifetime. Several questions remain regarding “severe disease risk” subtype participants; it is unclear whether these individuals had defects in lung growth and development that predisposed them to more severe disease earlier in life or if we simply did not capture a previously occurring period of disease activity and progression.
